# Evaluation of Flow Control Using PID versus Fuzzy Logic in an Electropneumatic Circuit for Pulmonary Ventilation Applications

**DOI:** 10.1101/2025.01.07.25320144

**Authors:** Alfredo Lescher, Lina González, Issa Griffith, Jay Molino, Asdrúal Rojas, Damián Quijano

**Affiliations:** Laboratorio de Investigación Experimental de Bioseñales, Centro I+D+i de Biotecnología, Energías Verdes y Cambio Climático, Universidad Especializada de las Américas (UDELAS), Albrook, Paseo de La Iguana 0843-014, Republic of Panama; Sistema Nacional de Investigación (SNI), SENACYT, Panama City, Republic of Panama

**Keywords:** Mechanical ventilation, PID control, Fuzzy logic control, Settling time, Stability analysis, Nyquist analysis, Transient oscillations

## Abstract

High-tech commercial mechanical ventilators are designed to provide a constant and accurate flow, thus ensuring the precision of ventilatory treatment. Therefore, this research aims to evaluate the flow control in a prototype electro-pneumatic unit for mechanical ventilator applications using a PID versus a Fuzzy Logic method.

Specifically, the intention is to determine if there is evidence of superior performance of fuzzy logic control over PID in this application. For this reason, the methodology is based on measurement and numerical analysis. The design of this research is quasi-experimental and conducted in a laboratory. Thus, samples of the data of the variables were taken at convenience under a pre-established scheme and conditions. In terms of the type of study, it is considered quantitative, experimental, and applied.

The main results show that according, the Bland-Altman analysis both controllers meet the accuracy limits of commercial equipment. However, the fuzzy logic controller presented better standard deviation and difference limits, demonstrating greater reliability in the accuracy of results. Additionally, the PID control demonstrated quicker response times with a shorter settling time (between 0.32 and 0.43 seconds) than in the fuzzy logic control (between 0.43 and 0.77 seconds); moreover, the fuzzy control presents an improvement in the volume of 900 mL, highlighting the efficiency of this controller in conditions of higher demand. The results show that both the PID and Fuzzy Logic controllers meet the stability conditions of the Jury Test. The system’s poles are within the unit circle of the Z plane, confirming the controllers’ stability. Also, the curve of the fuzzy control in the Nyquist analysis is farther away from the origin and from the point (−1, 0j) than the curve of the PID control, indicating better stability and a more robust system against variations or disturbances.

## 1. Introduction

Mechanical ventilators are vital respiratory assistance devices that integrate and control variables such as flow, volume, pressure, and time to administer normal breathing under positive pressure [1]. These ventilators require precise monitoring of pressure or volume and a stable flow response, necessitating control systems that ensure stability and exceptional performance. Among the most common control methods are Proportional-Integral-Derivative (PID) control and Fuzzy Logic control, each with advantages and limitations in biomedical applications. This study aims to determine the impact of these approaches in controlling respiratory flow.

The evolution of mechanical ventilation has been driven by the need to support critically ill patients with respiratory insufficiencies, such as pneumonia, which causes low oxygen levels, or conditions leading to high carbon dioxide levels, like chronic obstructive pulmonary disease (COPD) [2,3]. Early 20th-century techniques differ significantly from modern mechanical ventilation systems that employ positive pressure [4]. The 1990s marked a significant advancement with the introduction of microprocessor-based controls in ventilators, enhancing synchronization with patient needs [4]. Today, fourth-generation ventilation technology frequently applies feedback controls to ensure uniform pressure and flow waveforms, even under varying conditions [5].

Control systems in mechanical ventilation must ensure optimal performance and stability against disturbances and parameter variations [6]. Control engineering focuses on creating models of various physical systems and using these models to develop controllers that ensure feedback systems meet performance specifications such as stability, steady-state, transient behavior (including overshoot, settling time, rise time, and peak time), disturbance rejection, and robustness against modeling uncertainties [7].

Currently, manufacturers such as Hamilton, Dräger, and Mindray design state-of-the-art commercial ventilators to provide constant and precise flow, ensuring the accuracy of ventilatory treatment. These ventilators require detailed pressure monitoring and consistent flow delivery, often implementing PID control systems [8]. The PID control system is a straightforward regulatory mechanism that establishes a set point as the target in the control process, operating on closed-loop principles with negative feedback that continuously adjusts performance based on comparing the desired state and the system’s current state [9].

Despite the reliability of PID control in the industry, it has limitations, mainly when system dynamics are unstable. It is crucial that, during the ventilation process, the pressure is adjusted according to the required ventilation level to prevent lung damage [10]. While PID control excels in response times, fuzzy logic control surpasses it in several significant aspects [11]. Fuzzy logic control integrates common language into the control system design, eliminates the need for complex system modeling [12], and applies to nonlinear systems [13]. Additionally, it offers superior performance with shorter stabilization times, fewer transient oscillations, and reduced steady-state errors [14].

Introduced by Dr. Lotfi Zadeh in 1965, fuzzy logic can be mathematically represented to handle approximate reasoning and complex systems [13,15]. Various studies have demonstrated its effectiveness in dynamic systems, particularly in biomedical applications [15,16]. Fuzzy logic controllers, structured around fuzzification, a knowledge base of fuzzy rules, inference mechanisms, and defuzzification, provide a robust framework for managing complex control systems [17].

Recent studies have highlighted the equivalence and transformation between conventional PID controllers and fuzzy logic controllers (FLCs). For instance, Chao et al. (2017) [18] emphasized that a well-designed conventional PID controller could be transformed into an equivalent fuzzy logic controller by observing and defining the operating ranges of the input/output variables. This approach allows for leveraging the mature design techniques of PID controllers while benefiting from the nonlinearity of FLCs, which can yield more satisfactory system responses. This equivalence is crucial for understanding how FLCs can be tuned to outperform conventional PID controllers, especially in systems that exhibit nonlinear behavior.

Moreover, fuzzy logic controllers have been shown to handle nonlinearity more effectively than traditional PID controllers, particularly in complex dynamic systems. A study published in *PLOS ONE* (2023) [19] discussed the advantages of fuzzy logic in controlling systems with significant nonlinearity, where traditional PID controllers may struggle with delayed responses and overshoot issues. This supports the growing consensus that FLCs offer enhanced stability and adaptability in scenarios where system dynamics are not straightforward, thus justifying their use in biomedical applications such as mechanical ventilation.

This research evaluates a prototype electro-pneumatic unit for mechanical ventilators using PID and fuzzy logic methods to determine which control strategy offers superior performance. Specifically, it assesses whether fuzzy logic control can outperform PID control in terms of efficiency and stability in mechanical ventilation applications. This investigation is crucial in emerging and developing economies that need autonomous technology for medical devices, ensuring availability during unexpected events like epidemics or natural disasters [20,21].

By addressing these needs, the study aims to contribute to developing advanced medical equipment enhancing the capabilities of artificial ventilators to ensure patient safety and comfort during ventilation periods. The findings will also provide a foundation for future research and development of medical technologies, aligning with sustainable development goals and reducing inequalities in access to high-quality treatments with autonomous technologies.

## 2. Prototype

The purpose of the PID and Fuzzy Logic control in a prototype electro-pneumatic ventilator. The setup includes an ESP32 microcontroller, flow and pressure sensors, and control valves. PID control parameters were tuned for optimal response, while Fuzzy Logic control utilized specific rules and membership functions. Data collection involved measuring flow, volume, and pressure at a high sampling rate, followed by statistical analysis of performance metrics, including settling time, overshoot, and stability margins. Validation was performed against ISO 80601-2-12:2021 standards to ensure accuracy and reliability.

### 2.1 Test Equipment

The components used in the prototype setup include a precision test lung, model ACCU LUNG by Fluke, and a flow and pressure analyzer, model VT650 by Fluke. A digital oscilloscope, model 795-TBS1102C, from Tektronix and a digital multimeter, model 179, by Fluke, were utilized. The power supply used is model SPD3303X-E from Siglent Technologies. A medical-grade air compressor, Aridyne 3500, by Allied Medical LLC, was employed to provide the necessary airflow.

### 2.2 Electropneumatic unit and control

The prototype uses an ESP32 microcontroller for its high performance, dual-core processing, integrated Wi-Fi/Bluetooth, and extensive GPIO pins, essential for real-time data processing and control. The electronic circuit, illustrated in Figure 1 (left panel), uses the ESP32-DevKitC V4 development board with the ESP-WROOM-32 module from Espressif Systems [22] This module enables communication with the computer, which implements PID and fuzzy logic control codes. The microcontroller’s control signal regulates the state (open or closed) of the proportional and solenoid valves, which control the flow. A specifically designed electronic circuit is required to manage these valves effectively. The Electropneumatic Unit is structured into two main components: the inspiratory and expiratory sections, as illustrated in Figure 1 (right panel).

**Fig. 1.**
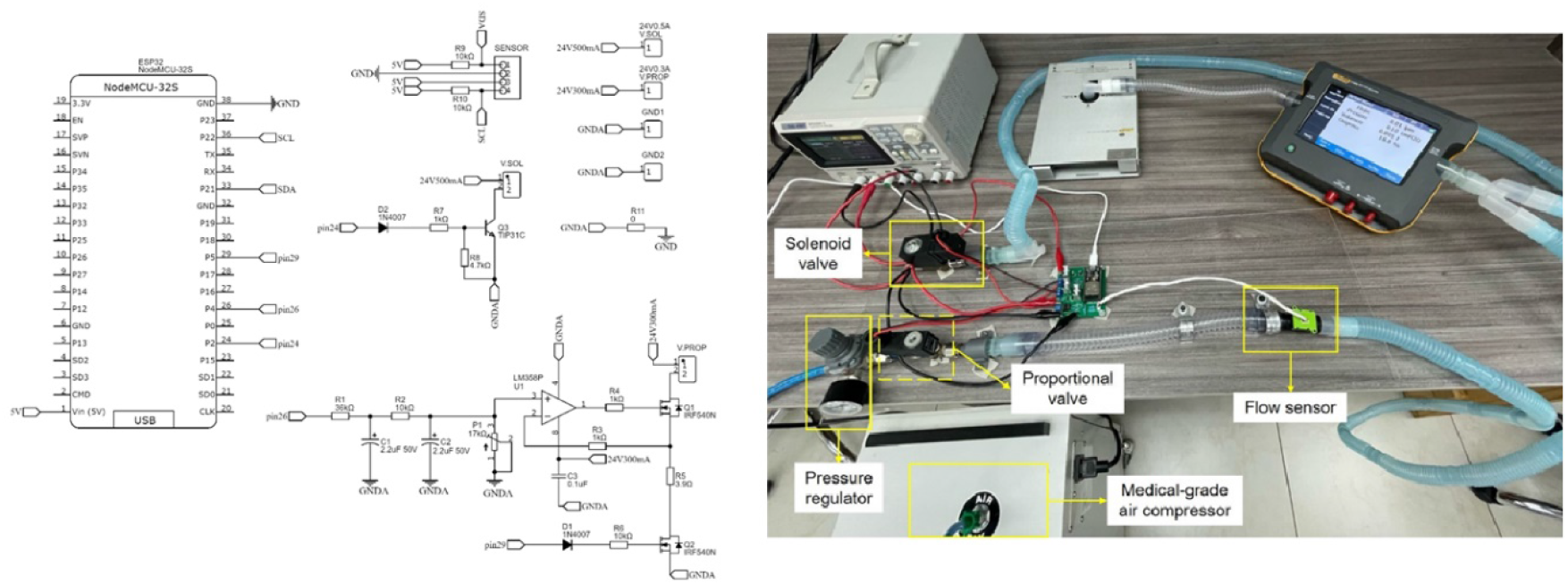
(Left panel) Schematic of the electronic circuit. (Right panel) Electropneumatic Unit.

The inspiratory section starts with Allied Medical LLC’s Aridyne 3500 medical-grade air compressor providing air at 50 psi [23]. A Series M micro regulator from Camozzi [24] adjusts this pressure to 30 psi. Following this, as detailed in its datasheet, a Series AP proportional valve from Camozzi [25] varies the flow proportionally with the applied current. PID and fuzzy logic control focus on this valve. The valve connects to a Sensirion SFM3000 flow sensor [26] that measures the flow and provides feedback to the control system before reaching the inspiratory branch of the patient circuit.

In contrast, the expiratory section includes a solenoid valve, Series CFB from Camozzi [27], which regulates the expulsion of air by alternating between its open and closed states, thus performing the expiratory function of the system. This solenoid valve connects on one end to the expiratory branch of the patient circuit. In contrast, the other end is left open to the outside air, facilitating the expulsion of air.

This setup integrates high-accuracy sensors, fast-response actuators, and the powerful ESP32 to create a robust platform for evaluating PID and Fuzzy Logic control strategies in mechanical ventilation.

### 2.3 Patient Circuit

The patient circuit comprises the inspiratory and expiratory branches connected to the patient. In the context of the laboratory tests conducted, a test lung was used instead of human patients. This methodology allows for verifying the accuracy of inspiratory flow control. The circuit is connected to the Fluke VT650 Flow Analyzer for these experimental tests, chosen for its high precision, reliability, and portability in verifying respiratory medical equipment [28]. The ACCU LUNG model, also from Fluke, was used as the test lung, and it was noted for its ability to adjust to three different types of compliance and resistance conditions in the airway, thereby simulating the possible conditions of an actual patient [29].

### 2.4 Development of the PID Control Algorithm

The PID control algorithm was implemented using the Python library “simple_pid,” which is user-friendly and requires no external dependencies. A PID object is created to define the constants *K*_*p*_ = 5.7, *K*_*i*_ = 87.3 and *K*_*d*_ = 0.05, and the setpoint. The PID control algorithm settings include output limits between 540 and 1023, with a sampling time set at 0.5 milliseconds. The system operates in “Auto” mode, enabling automatic calculation of new control values. Within the main loop, the input (measured variable) and the controller’s output (the variable to adjust) are defined. The PID controller then calculates the feedback signal based on the error between the setpoint and the current value, and this signal is subsequently applied to the system to achieve the desired control.

### 2.5 Development of the Fuzzy Logic Control Algorithm

The development of fuzzy logic control began with analyzing the system to create universes of discourse, membership functions, linguistic variables, and control rules. The controller was structured with two inputs, one output, and 12 rules (Table 1).

**Table 1.**
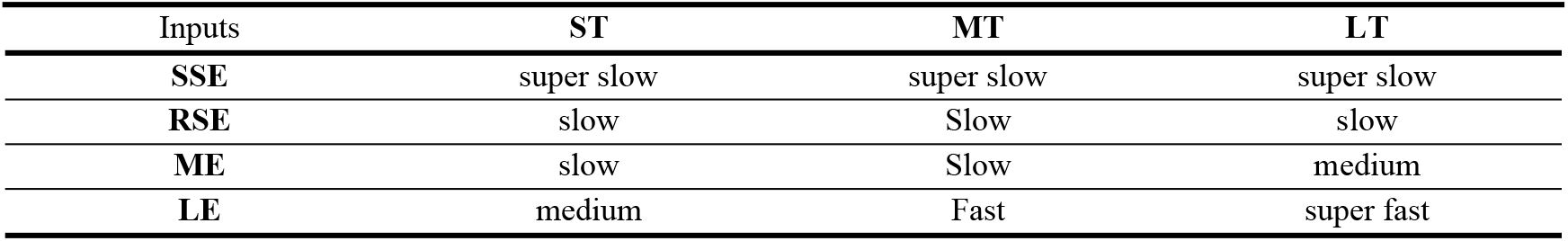
Fuzzy Control Rules.

The fuzzy logic control system was developed with specific inputs and outputs. The first input is the relative flow error, which ranges from 0 to 100 and is calculated by comparing the sensor-measured flow with the target flow. This input has four trapezoidal membership functions: super small (0-8%), relatively small (3-30%), medium (15-50%), and large (40-100%). The second input is the target flow, which ranges from 0 to 35 l/min and has three trapezoidal membership functions: small (0-21 L/min), medium (20-30 L/min), and large (26-35 L/min).

The output of the system is a factor added to the PWM duty cycle, defined by five trapezoidal membership functions: super slow (0-0.08), slow (0.07-0.4), medium (0.3-2), fast (1.8-3.5), and super fast (3-4). The fuzzy logic controller uses 12 rules to determine the output based on the inputs. For example, if the target flow is small and the relative error is negligible, the output is super slow; if the target flow is large and the relative error is large, the output is super fast.

The fuzzy control was implemented using the “skfuzzy” Python library and a Lookup Table (LUT) for efficiency, avoiding extensive real-time calculations. Due to the ESP32’s limited memory, the LUT was stored and executed on a PC via serial communication. Three key files were created for the implementation: “fuzzy_generador_matriz.py” defines membership functions and control rules to generate LUTs; “fuzzy_comunicacion.py” is the main program for receiving sensor data, performing LUT lookups, and sending PWM values to the ESP32; and “boot.py” manages respiratory timing logic and runs on the ESP32 at 240 MHz. The serial communication operated at 115200 baud with a PC using an Intel Core i7 processor, overcoming previous speed limitations. This approach demonstrated functionality and effectiveness, allowing for control adjustments and testing.

### 2.6 Ethical Statement

This study did not require ethics approval because it was conducted using a prototype electro-pneumatic ventilator tested exclusively with simulators and not with human subjects. The experimental setup employed a precision test lung (ACCU LUNG by Fluke) and a flow and pressure analyzer (VT650 by Fluke) to validate the system’s performance. These devices are widely recognized for their reliability in simulating human respiratory conditions under controlled laboratory environments. As no human participants or biological samples were involved, ethical approval was not necessary for this research

## 3. Results

The analysis covers several performance metrics, including settling time, overshoot, volume accuracy, stability margin, and transient oscillations.

### 3.1 Conformity Evaluation

The conformity evaluation was conducted using the electro-pneumatic test bench, comparing its measurements with the Fluke VT650 flow analyzer under both PID and Fuzzy Logic control. A Bland-Altman analysis revealed mean differences of -0.122 L/min for PID and -0.193 L/min for Fuzzy Logic control (Table 2).

**Table 2.** Bland-Altman Analysis Results.

| Control | Parameter | Value | S.E. | Inferior | Superior |
| --- | --- | --- | --- | --- | --- |
| PID | Mean Difference | -0.122 | 0.059 | -0.238 | -0.005 |
|  | Lower Limit | -1.269 | 0.100 | -1.468 | -1.070 |
|  | Upper Limit | 1.025 | 0.100 | 0.826 | 1.225 |
| Fuzzy Logic | Mean Difference | -0.193 | 0.055 | -0.302 | -0.084 |
|  | Lower Limit | -1.263 | 0.094 | -1.449 | -1.076 |
|  | Upper Limit | 0.876 | 0.049 | 0.690 | 1.063 |

The standard error of the mean (S.E.) was 0.059 for PID and 0.055 for Fuzzy Logic control. The limits of variability ranged from -1.2688 to 1.0255 L/min for PID and from -1.2627 to 0.8765 L/min for Fuzzy Logic control.

The mean differences observed for both controls compared to the VT650 analyzer were not significant, aligning with the precision of recognized commercial equipment [30], [31], [32]. The experimental readings were precise, suggesting robustness in the control system’s ability to replicate measurements under controlled conditions.

The Bland-Altman graph (Figure 2) visualized these differences against the average measurements of the control and analyzer. The distribution of disagreements showed no clear pattern of proportional bias, indicating that discrepancies did not systematically increase or decrease with the measured flow levels.

**Fig. 2.**
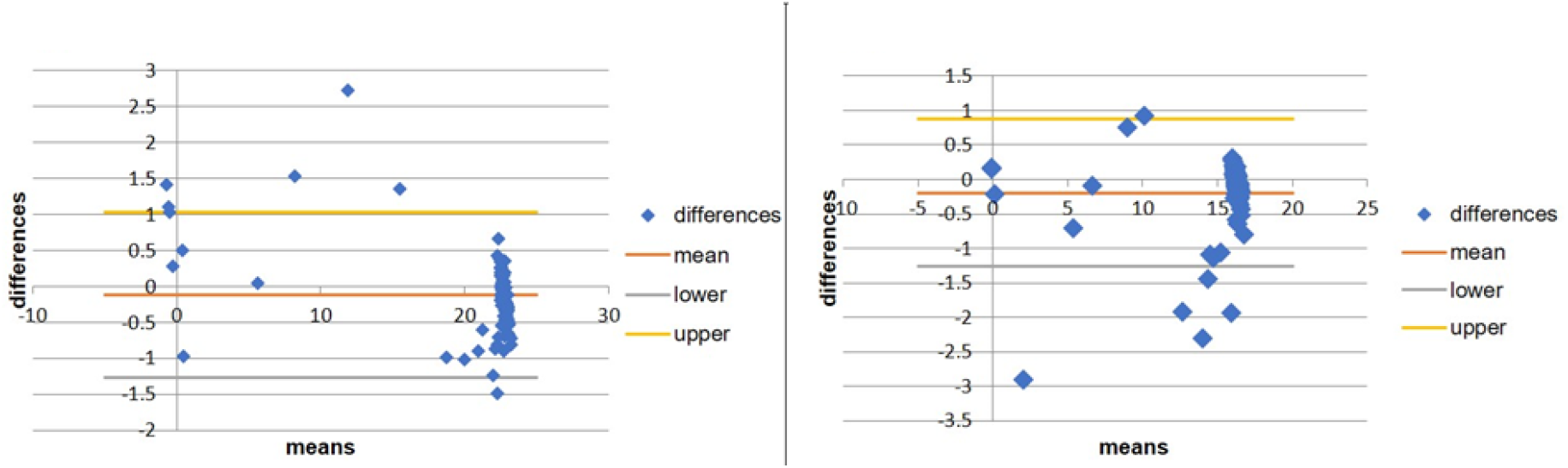
Bland-Altman Analysis. (Left Panel). PID Control. (Right Panel) Fuzzy Logic Control.

The Bland-Altman analysis for each volume test with both controllers demonstrated notable similarity and consistency with previous results. Both controls fell within the precision limits of recognized commercial equipment. Still, the Fuzzy Logic control showed better standard deviation and limits of differences, indicating higher reliability in reproducing consistent and predictable flow results, which is crucial in managing critical patients where ventilator stability is essential.

The visual inspection of the Bland-Altman graphs (Figure 3) showed that most differences for Fuzzy Logic control were closely clustered around the mean line and within the limits of agreement, indicating higher precision and consistency. The mean difference line for Fuzzy Logic control was closer to zero, suggesting a less average underestimation of flow compared to the Fluke analyzer. Additionally, the differences beyond the limits were less dispersed, indicating fewer and less pronounced extreme errors than PID control.

### 3.2 Repeatability

The Fisher-Snedecor statistical test was applied to evaluate repeatability, calculating the required sample size using the formula for a finite sample (Equation 1) [33].

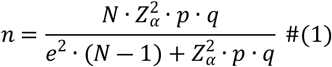

The sample size calculation is based on several key parameters. Here, n represents the sample size, while N denotes the population size. The statistical parameter Z corresponds to the confidence level, and e is the maximum acceptable error. The probability of success is indicated by p, with q representing the probability of failure, defined as (1 ― *p*). For the electro-pneumatic test bench, with a population of 4000 records, a 95% confidence level (Z=1.96Z = 1.96Z=1.96) and a 5% maximum acceptable error were used, resulting in a calculated sample size of 71.70. Similarly, for the analyzer, which had a population of 2000 records, the sample size was determined to be 70.40.

Stable state data were selected for the Fisher test with a significance level of 0.05, applied to each controller and volume test. Results are shown in Tables 3 and 4.

**Table 3.** Fisher Test for PID Controller.

| Volume (mL) | Test Bench Variance | Analyzer Variance | Test Bench Std Dev | Analyzer Std Dev | F | P(F<=f) | Critical F |
| --- | --- | --- | --- | --- | --- | --- | --- |
| 500 | 0.003 | 0.002 | 0.054 | 0.050 | 1.191 | 0.233 | 1.487 |
| 700 | 0.003 | 0.004 | 0.056 | 0.063 | 1.280 | 0.152 | 1.485 |
| 900 | 0.004 | 0.004 | 0.064 | 0.065 | 1.033 | 0.446 | 1.485 |

**Table 4.** Fisher Test for Fuzzy Logic Controller.

| Volume (mL) | Test Bench Variance | Analyzer Variance | Test Bench Std Dev | Analyzer Std Dev | F | P(F<=f) | Critical F |
| --- | --- | --- | --- | --- | --- | --- | --- |
| 500 | 0.002 | 0.002 | 0.050 | 0.048 | 1.093 | 0.355 | 1.487 |
| 700 | 0.003 | 0.004 | 0.052 | 0.063 | 1.469 | 0.055 | 1.485 |
| 900 | 0.003 | 0.004 | 0.056 | 0.059 | 1.113 | 0.327 | 1.485 |

The F-test results compare the means’ hypotheses:

- *Null Hypothesis (H0): No statistically significant difference between group variances*.
- *Alternative Hypothesis (H1): Statistically significant difference between group variances*.

Hypotheses are accepted or rejected by comparing F values to the critical F value. If F < critical F, the null hypothesis is rejected. Additionally, P values were compared to the significance level (P ≤ 0.05) for validation.

Tables 3 and 4 analysis shows consistent results supporting the null hypothesis. F values for the PID controller (500 ml, 700 ml, 900 ml) were 1.191, 1.280, and 1.033, all below critical F values. For the Fuzzy Logic controller, F values (500 ml, 700 ml, 900 ml) were 1.093, 1.469, and 1.113; all values are also below critical F value. These results are enough to declare statistical significance for both controllers.

Besides, P values for Fuzzy Logic controller tests were consistently above 0.05, reinforcing the null hypothesis. These results indicate robust control systems maintaining volume delivery homogeneity under test conditions, suggesting high repeatability.

### 3.3 Settling time

The settling time refers to the period required for the system output to remain within a specified range between 2% and 5% of the desired final steady-state value [33]. For this analysis, we define the settling time as when the output remains within ±2% of the final value.

The PID controller showed exceptional settling time performance, with average times of 0.43s for 500 mL, decreasing to 0.35s for 700 mL and 0.32s for 900 mL. It also indicates a fast response, which is crucial in clinical situations requiring precise adjustments.

In contrast, the Fuzzy Logic controller presented longer times, with 0.77s for 500 mL, improving to 0.43s for 700 mL and 0.58s for 900 mL.

### 3.4 Stability

#### Jury Analysis

Stability was evaluated using Jury analysis of the transfer functions for each control, obtained with Matlab’s System Identification Toolbox. The resulting transfer function for the Fuzzy Logic control is presented in Equation 2. Equation 3 shows the transfer function obtained for the PID control.

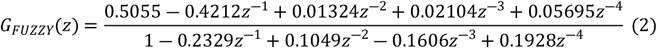

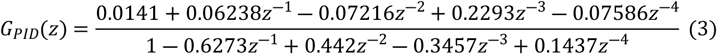

The results indicate that both the PID and Fuzzy Logic controllers meet the four conditions established by the Jury Test [35]. Once all the stability conditions are satisfied, it is concluded that the given characteristic equations are stable. Furthermore, when graphically verifying the system’s poles, they all appear within the unit circle of the Z plane. This confirms that the stability criteria for the discrete mode controllers of the presented design are met.

#### Nyquist Analysis

Nyquist plots (Figure 4) show the relationship between the real and imaginary parts of the system’s frequency response. Both controls do not encircle the critical point (−1, 0j), indicating stability [35].

**Fig. 4.**
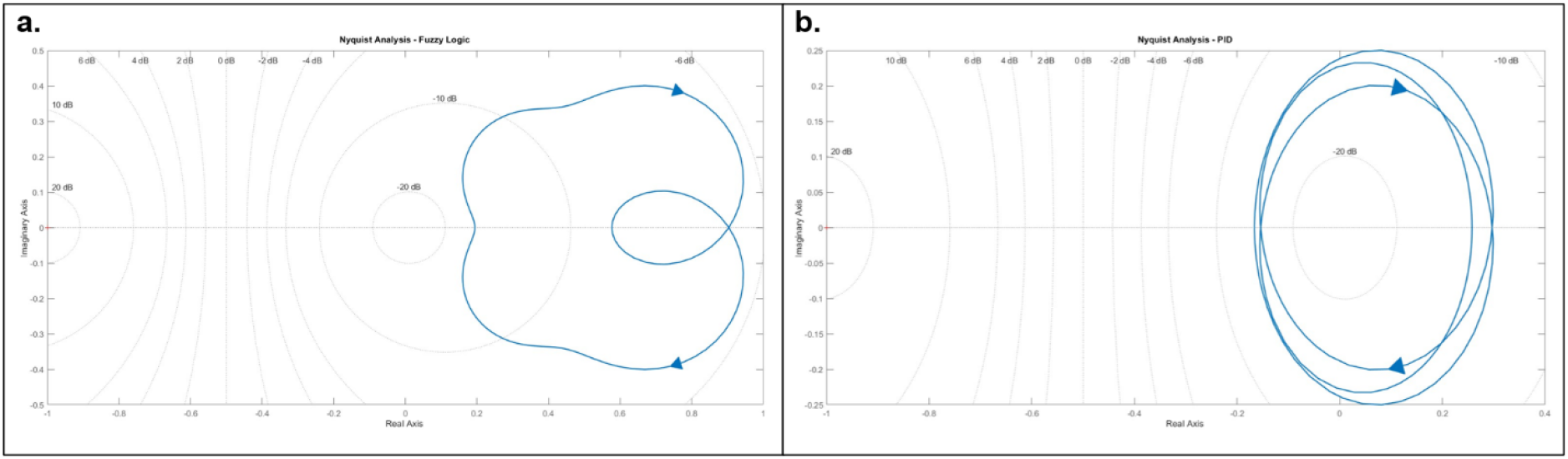
Nyquist diagrams: A. Fuzzy Logic Control. B. PID Control

The fuzzy logic control plot (Figure 4. a) is farther from the origin and the critical point than PID control (Figure 4.b), indicating better stability and robustness against variations. The multiple loops in the PID control plot suggest pronounced frequency response peaks, indicating sensitivity at specific frequencies. The fuzzy logic control plot indicates smoother, less resonant behavior, which is beneficial for robustness against operational variations and disturbances.

Gain and phase margins were also evaluated. Fuzzy logic control showed infinite gain and phase margins, indicating tolerance to any gain or phase shift before instability. PID control had a gain margin of 15.58 dB, indicating some tolerance to gain before instability.

## 4. Discussion

The experimental evaluation of the fuzzy logic and PID controllers for the electro-pneumatic ventilator system demonstrates several key findings that highlight the advantages of the fuzzy logic controller. The results show that both systems are within the accuracy margins of commercial equipment. Still, Fuzzy Logic, with lower variability in measurements according to Bland-Altman analysis, appears more accurate and consistent. Regarding repeatability, when comparing the results of both controllers with the established target flows, it is perceived that both Fuzzy Logic and PID have a high capacity to reach the target flows with minimal deviations.

Although PID achieves faster settling times, Fuzzy Logic fits better at higher volumes, showing better adaptability, which is crucial in dynamic clinical environments. Stability analyses, including Nyquist plots and Jury analysis, indicate that both systems are stable. However, the Fuzzy Logic controller exhibits a smoother and less resonant frequency response, suggesting greater robustness to variations, which is beneficial in complex environments.

Further development and refinement of fuzzy logic algorithms are essential. Optimizing parameters and incorporating machine learning techniques could enhance the adaptability and precision of the controller, allowing it to adjust dynamically to real-time data. Controlling flow in neonatal patients presents unique challenges due to the low volumes and high frequencies involved. Future studies should focus on this scenario to ensure that the fuzzy logic controller can handle these demanding conditions effectively. Both controllers’ algorithms should be optimized, especially for high-flow situations where the PID controller has shown better responsiveness. Enhancing the fuzzy logic controller’s performance in these conditions could further establish its superiority.

## Data Availability

All relevant data are within the manuscript and its Supporting Information files.

## 5. Authors contributions

**AL, LG**, and **IG** equally worked on the conceptualization, data curation, formal analysis, funding acquisition, investigation, methodology, project administration, resources, software, supervision, validation, visualization, Writing – review & editing **AR, JM, DQ** equally worked on resources, software, supervision, validation, visualization, Writing – original draft, Writing – review & editing.

## 7. Acknowledgments

The authors would like to acknowledge the support of SENACYT via grant No. APY-NI2022-13.

## 8. Conclusion

This study has demonstrated the superior performance of Fuzzy Logic control over traditional PID control in the context of mechanical ventilation systems, particularly in terms of stability and robustness. The data analysis clearly shows that Fuzzy Logic control provides enhanced adaptability and precision, making it a more effective solution for managing mechanical ventilation’s complex and dynamic requirements. These findings are especially relevant in scenarios where patient-specific adjustments and nonlinearities in system behavior are critical. For practitioners, the results of this study suggest that integrating Fuzzy Logic control into mechanical ventilation systems could significantly improve patient outcomes by offering more consistent and reliable respiratory support. Practitioners should consider the adoption of Fuzzy Logic controllers in environments where precise control of ventilation parameters is essential, such as in intensive care units or during critical surgical procedures. For policymakers, the research underscores the importance of supporting the development and implementation of advanced control technologies in medical devices. Policies encouraging the integration of Fuzzy Logic control in medical equipment could improve healthcare delivery, particularly in emerging and developing economies where access to high-quality medical technology is often limited. Additionally, establishing standards and guidelines for using Fuzzy Logic control in medical devices could further enhance patient safety and care quality.

## References

[1] Walter JM, Corbridge TC, Singer BD. Invasive Mechanical Ventilation. Southern Medical Journal 2018;111:746–53. 10.14423/SMJ.0000000000000905.

[2] Major VJ, Chiew YS, Shaw GM, Chase JG. Biomedical engineer’s guide to the clinical aspects of intensive care mechanical ventilation. Biomedical Engineering Online 2018;17:169. 10.1186/s12938-018-0599-9.

[3] Walter K. Mechanical Ventilation. Journal of the American Medical Association (JAMA) 2021;326:1452.

[4] Soto G G. Ventilación Mecánica: Una Breve Historia. Neumología Pediátrica 2016;11:151–4. 10.51451/np.v11i4.288.

[5] Hess DR, Kacmarek RM. Principles of Mechanical Ventilation. Essentials of Mechanical Ventilation. 4th ed., McGraw Hill; 2018, p. 47–8.

[6] López CP. MATLAB Control Systems Engineering. 1st ed. Berkeley, CA: Apress Berkeley, CA; 2014. 10.1007/978-1-4842-0289-0.

[7] Dorf RC, Bishop RH. Introduction to Control Systems. Modern Control System. 14th ed., Pearson; 2021, p. 30.

[8] Hunnekens B, Kamps S, Van De Wouw N. Variable-Gain Control for Respiratory Systems. IEEE Transactions on Control Systems Technology 2020;28:163–71. 10.1109/TCST.2018.2871002.

[9] Baltieri M, Buckley C. PID Control as a Process of Active Inference with Linear Generative Models. Entropy 2019;21:257. 10.3390/e21030257.

[10] Mehedi IM, Shah HSM, Al-Saggaf UM, Mansouri R, Bettayeb M. Fuzzy PID Control for Respiratory Systems. J Healthc Eng 2021;2021:1–6. 10.1155/2021/7118711.

[11] Kiyak E, Gol G. A comparison of fuzzy logic and PID controller for a single-axis solar tracking system. Renew Wind Water Sol 2016;3:7. 10.1186/s40807-016-0023-7.

[12] Montoya Giraldo OD, Valenzuela Hernández JG, Giraldo Buitrago D. Lógica Difusa Aplicada al Control Local del Péndulo Invertido con Rueda de Reacción. Scientia Et Technica 2013;18:623–32.

[13] Matute Clavier A, Bernal Suárez WF. Técnicas de lógica difusa en ingeniería de control. Ciencia, Innovación y Tecnología (RCIYT) 2017;3:125–34.

[14] Ontiveros JJ, Ávalos CD, Loza F, Galán ND, Rubio GJ. Evaluation and Design of Power Controller of Two-Axis Solar Tracking by PID and FL for a Photovoltaic Module. International Journal of Photoenergy 2020;2020:1–13. 10.1155/2020/8813732.

[15] El Adawy MI, El-Garhy AM, Sawafta FO. Design of Fuzzy Controller for Supplying Oxygen in Sub-acute Respiratory Illnesses. International Journal of Computer Science Issues (IJCSI) 2012;9:192–206.

[16] Ospino Castro A, Robles Algarín C, Duran Pabón A. Diseño de un sistema médico asistencial de autorregulación de oxígeno por monitoreo no invasivo, basado en lógica difusa. Prospectiva 2014;12:57. 10.15665/rp.v12i2.289.

[17] Danapalasingam KA. Robust Fuzzy Logic Stabilization with Disturbance Elimination. The Scientific World Journal 2014;2014:1–7. 10.1155/2014/171597.

[18] C.-T. Chao, N. Sutarna, J.-S. Chiou and C.-J. Wang, “Equivalence between Fuzzy PID Controllers and Conventional PID Controllers,” Applied Sciences, pp. 1-12, 2017.

[19] A. Daraz, A. Basit and G. Zhang, “Performance analysis of PID controller and fuzzy logic controller for DC-DC boostconverter,” PLoS ONE, vol. 18, no. 10, pp. 1–18, 2023.

[20] Yousfi Allagui N, Salem FA, Aljuaid AM. Artificial Fuzzy-PID Gain Scheduling Algorithm Design for Motion Control in Differential Drive Mobile Robotic Platforms. Computational Intelligence and Neuroscience 2021;2021:1–13. 10.1155/2021/5542888.

[21] Gereffi G. What does the COVID-19 pandemic teach us about global value chains? The case of medical supplies. Journal of International Business Policy 2020;3:287–301. 10.1057/s42214-020-00062-w.

[22] Espressif Systems. ESP32-DevKitC V4 Getting Started Guide. Espressif 2024. https://docs.espressif.com/projects/esp-idf/en/release-v4.0/hw-reference/get-started-devkitc.html (accessed February 21, 2024).

[23] Allied Medical LLC. Timeter® Aridyne™ Compresores de aire. Allied Medical LLC 2024. https://alliedmedicalllc.com/es/product/timeter-aridyne-air-compressors/ (accessed February 25, 2024).

[24] Camozzi. Catálogo Resumen. Camozzi 2021. https://camozzi.shop/pdf/Catalogo_Resumido.pdf (accessed February 25, 2024).

[25] Camozzi. Catálogo: Tecnología Proporcional. Camozzi 2019. https://ar.automation.camozzi.com/kdocs/2068439/06_tecnologa_proporcional_es-6-lowplus.pdf (accessed February 25, 2024).

[26] Sensirion. Datasheet SFM3000. Sensirion 2016. https://sensirion.com/media/documents/BA2F2B48/616681DC/Sensirion_Mass_Flow_Meters_SFM3000_Datasheet.pdf (accessed February 25, 2024).

[27] Camozzi. Valves and Solenoid Valves. Camozzi 2023. https://media.camozzi.com/pdf/ENG.4.1.31.pdf (accessed February 25, 2024).

[28] Fluke. Fluke Biomedical Medical Gas Flow Analyzers. Fluke Biomedical 2024. https://acortar.link/duVYms (accessed February 25, 2024).

[29] Fluke. ACCU LUNG Precision Test Lung. Fluke Biomedical 2024. https://www.flukebiomedical.com/products/biomedical-test-equipment/gas-flow-analyzers-ventilator-testers/accu-lung-precision-test-lung (accessed February 25, 2024).

[30] Dräger. Instructions for use Savina 300 2020:204.

[31] Hamilton Medical. Manual del operador HAMILTON-C6 2021:360.

[32] Mindray. SV600 Ventilator Operator’s Manual 2018:240.

[33] Martínez Bencardino C. Distribuciones muestrales. Muestreo aleatorio. Estadística y muestreo. 14th ed., Ecoe Ediciones; 2019, p. 303– 6.

[34] Raol JR, Ayyagari R. Discrete Time Control System. Control Systems. 1st ed., CRC Press; 2019, p. 300. 10.1201/9781351170802.

[35] Fadali MS, Visioli A. Stability of Digital Control Systems. In: Academic Press, editor. Digital Control Engineering: Analysis and Design. 3rd ed., Elsevier Inc.; 2020, p. 117–37.

